# Integrating feature importance techniques and causal inference to enhance early detection of heart disease

**DOI:** 10.1101/2024.08.11.24311833

**Authors:** Atousa Arzanipour

## Abstract

Heart disease remains a leading cause of mortality worldwide, necessitating robust methods for its early detection and intervention. This study employs a comprehensive approach to identify and analyze critical features contributing to heart disease. Using a dataset of 270 patients, three well-known feature importance techniques—Boruta, Information Gain, and Lasso Regression—are applied to determine the top five features for heart disease detection. Following the identification of these key features, the g-computation method, a causal inference technique, is utilized to explore the causal relationships between these features and the presence of heart disease. The findings provide valuable insights into not only the features that are highly correlated with chronic heart disease but also those that have a direct causal impact on the classification of patients. This integrated approach enhances the understanding of heart disease etiology and can inform more effective diagnostic and therapeutic strategies.

## Introduction

Chronic diseases are long-term health conditions that persist over time and require ongoing medical management. Examples include cardiovascular diseases (e.g., heart disease), diabetes, chronic respiratory diseases, and various types of cancer. These conditions often necessitate continuous treatment to manage symptoms and prevent complications. According to CDC yearly reports, six out of ten Americans live with at least one chronic disease, making these diseases the leading causes of death and disability in the United States. They are also the primary drivers of healthcare costs [11]. Addressing chronic diseases should be a priority for the United States, as reports estimate that by 2030, 171 million adults will have one or more chronic conditions [5].

In the case of heart disease prevention, various measures have been undertaken by different groups. On an individual level, many people are making more informed decisions and adopting healthier lifestyle habits. On a community level, wellness workshops and initiatives in schools and workplaces aim to educate people about the risks of heart disease and methods to reduce its occurrence within their communities. Additionally, governments have implemented broader strategies and regulations, such as controlling the availability of harmful substances (e.g., tobacco, alcohol) and providing national access to healthcare services for preventive care and affordable medications. Substantial funding has also been allocated by governments to study the causes of heart disease and its development.

ne area of research that has gained significant attention in recent years is the use of artificial intelligence, specifically machine learning models, in detecting heart disease. These models predict the presence of heart disease based on information from various patients, known as predictor variables in the literature. While these methods can assist healthcare providers in making more informed decisions, they often do not explore the deeper, sometimes confounding relationships between predictor variables and heart disease detection. Recently, there has been a shift toward causal analysis, which examines the causal effects between variables beyond mere correlation. For example, research has shown that an increase in the consumption of organic food in Seattle was correlated with a rise in autism cases. This high correlation between autism and organic food consumption is suspicious, and causal inference techniques have demonstrated that there is no causal relationship between the two—only a coincidental correlation. Therefore, causal analysis can be effectively applied in the healthcare industry to identify strong causal relationships between factors affecting heart disease detection.

In the upcoming sections, we will review the existing literature and identify the research gap. The Methods and Materials section will provide insights into the dataset and its preprocessing methods. It will also outline the computational steps of the causal inference technique, g-computation,used to identify the relationship between key predictor variables and heart disease detection in this dataset. Subsequently, the Results and Discussion section will elaborate on the experiment’s findings and explore potential future research paths.

## Literature Review

Feature selection in machine learning refers to the process of analyzing all the predictor variables and their effects on the response variable to achieve more accurate predictions in a faster and more efficient manner. When working with a dataset, not all features have an influence on the outcome, so there are many feature selection methods designed to identify and remove irrelevant features [1–4]. Essentially, feature selection involves choosing a method to search for the best features, defining evaluation criteria, and determining a stopping point [6]. Feature selection has been used in numerous studies to identify the most influential factors in heart disease detection, leading to more efficient predictions. Pal et al. employed feature selection methods, such as Extra Trees, in conjunction with well-known machine learning techniques like Support Vector Machines and Bagging methods. The results demonstrated an increase in accuracy.

In the realm of literature, extensive research has explored machine learning algorithms such as tree-based models, neural networks, and basic regression models. These methods aim to predict specific outcomes based on existing predictor variables. This trend is evident in chronic disease management, where input data like blood pressure, family history, and medication usage are used to assess whether a patient is at high risk for one or more chronic diseases [15–18]. However, this evaluation might not fully capture the intricate and subtle interconnections among various predictor variables and the eventual outcome. As a result, there is growing interest in using causal inference techniques to achieve more accurate predictions.

A significant body of literature has concentrated on exploring causal inference techniques to enhance the management of chronic diseases, with researchers delving into various aspects of this field [23–25]. Some studies have investigated the compounding effects of chronic diseases and their impact on hospital re-admissions, as demonstrated by Casucci et al. [5]. In parallel, other scholars have conducted causal inference analyses specifically tailored to chronic kidney diseases [19]. Notably, John Lee and colleagues have contributed to this area by focusing on feature selection techniques in their study titled “Chronic Disease Outcome Prediction Using a Causal Inference Technique.” They utilized data from electronic health records (EHR) to refine their approach [20].

Various methods exist for analyzing the causal relationship between two variables, with g-computation standing out as a crucial approach. Also referred to as the G-formula, g-computation is a statistical technique used in causal inference to determine the impact of an exposure or treatment on an outcome, even in the presence of confounding variables. This method is particularly valuable in observational studies where conducting randomized controlled trials is either not possible or ethically permissible. In the realm of epidemiology, there is a wealth of research dedicated to exploring g-computation and its applications in understanding causality [21]. In a notable study, Snowden et al. investigated chronic asthma by conducting a synthetic data study to effectively implement g-computation [8]. This underscores the method’s versatility and importance in unraveling complex causal relationships in real-world scenarios.

Correlation does not imply causation, and to our knowledge, there has been no prior research addressing this by extracting the most important and highly correlated features using feature selection methods and then thoroughly investigating the cause-and-effect relationship between key health factors in patients diagnosed with heart disease. These factors include cholesterol levels, sex, blood flow in veins, and the number of blocked veins, as well as their potential impact on the increased risk of heart disease. We will utilize the G-estimation technique to explore these relationships, and if a causal relationship is established, we will provide guidelines that can be used by individuals, healthcare providers, and policymakers to reduce the impact of this chronic health issue. In the next section, we will detail the dataset we intend to use for the G-estimation technique and describe the method itself.

## 1 Materials and Methods

In this study, we analyze a dataset of 270 patient records to identify the most important features contributing to the detection of heart disease. We then conduct a causal analysis to better understand the underlying relationships among these selected features. Using the well-established causal inference technique of g-computation, we investigate whether there is a causal relationship between these factors and the detection of heart disease, aiming to establish a robust causal connection rather than merely identifying correlations.

### 1.1 Materials

The name of the dataset used in this research is *Heart Disease Prediction*, and it can be accessed as a *csv* file. This dataset is shared on the Kaggle website and can be accessed using the following link.

This dataset encompasses 270 individual case studies, constituting 270 rows in total. These cases are categorized based on the presence (coded as 1) or absence (coded as 0) of heart disease, as determined through cardiac categorizations. Each patient is characterized by 13 distinct predictive factors, such as age, gender, chest pain type, blood pressure readings, cholesterol levels, and more. Table 1 provides a detailed overview of the dataset’s column names and their descriptions. Notably, this dataset is free of missing values, eliminating the need for any imputation methods.

**Table 1.**
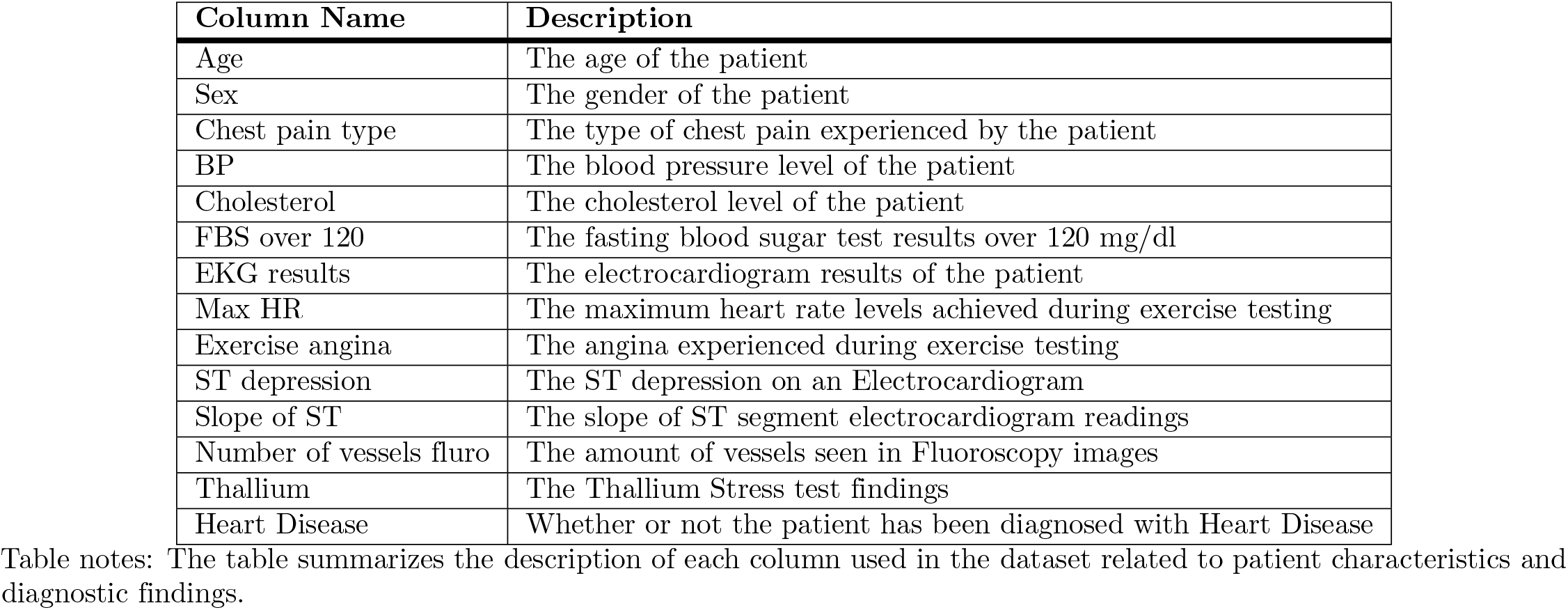
Description of Columns.

| Column Name | Description |
| --- | --- |
| Age | The age of the patient |
| Sex | The gender of the patient |
| Chest pain type | The type of chest pain experienced by the patient |
| BP | The blood pressure level of the patient |
| Cholesterol | The cholesterol level of the patient |
| FBS over 120 | The fasting blood sugar test results over 120 mg/dl |
| EKG results | The electrocardiogram results of the patient |
| Max HR | The maximum heart rate levels achieved during exercise testing |
| Exercise angina | The angina experienced during exercise testing |
| ST depression | The ST depression on an Electrocardiogram |
| Slope of ST | The slope of ST segment electrocardiogram readings |
| Number of vessels fluro | The amount of vessels seen in Fluoroscopy images |
| Thallium | The Thallium Stress test findings |
| Heart Disease | Whether or not the patient has been diagnosed with Heart Disease |

Table notes: The table summarizes the description of each column used in the dataset related to patient characteristics and diagnostic findings.

**Table 2.** Prediction Accuracy Using Selected Features.

| Feature Selection Method | Logistic Regression | Random Forest |
| --- | --- | --- |
| Full Feature Set | 81.48 | 79.01 |
| Boruta | 80.24 | 79.01 |
| Lasso Regression | 80.24 | 75.30 |
| Information Gain | 71.60 | 67.90 |

Table notes: The table shows the prediction accuracy of different models using selected feature sets.

**Table 3.** Results of the Analysis.

| Original Estimates |  |  |  | logOR Estimates |  |  |  |
| --- | --- | --- | --- | --- | --- | --- | --- |
|  | Estimate | ci.lower | ci.upper |  | Estimate | ci.lower | ci.upper |
| Thalium | 0.4495 | 0.0613 | 0.3292 | Thalium | 1.9382 | 0.3141 | 1.3225 |
| Sex | 0.1542 | 0.0612 | 0.0342 | Sex | 0.6243 | 0.2584 | 0.1178 |
| Vessel | 0.2790 | 0.1051 | 0.0736 | Vessel | 1.4234 | 3.6496 | -5.7296 |
| Cholesterol | 0.1623 | 0.0492 | 0.0659 | Cholesterol | 0.6694 | 0.2098 | 0.2581 |

Note: Results include original estimates and logOR estimates.

### 1.2 Methods

As shown in Table 1, this dataset includes 13 independent predictor variables and one dependent response variable, which is *Heart Disease*. However, not all of the independent predictor variables may be equally important in predicting whether a patient is diagnosed with heart disease. The literature includes a wealth of research dedicated to developing methods for identifying the most relevant features for predicting a response variable. This research employs three prominent methods of feature selection: The first is a wrapper around the Random Forest classification algorithm implemented in a package named *Boruta* in *R*, which we refer to as the *Boruta* method for brevity [22]. The other two methods are information gain and Lasso regression.

In all these feature selection methods, the 13 features are ranked based on their importance. To ensure that the features selected by these three methods work effectively, an experiment is conducted. The entire dataset, consisting of 270 rows and 13 predictor variables, is used in two well-known classification models: Logistic Regression and Random Forest. Seventy percent of the rows are utilized as the training dataset, while the remaining 30% serve as the test dataset to evaluate the models’ performance on unseen data. Accuracy is measured based on predictions made on this unseen data.

Next, for each feature selection method, the 5 most important features are identified. The same two classification models, Logistic Regression and Random Forest, are then applied to the dataset containing these 5 features and one response variable, using the same 70/30% train/test split. The results for accuracy are summarized in Table **??**.

As evident from the results, despite excluding the other predictor variables, models using only the five most important features achieved high prediction accuracy compared to models using all features.

Figures 1, 2, and 3, illustrates the importance of all features across three different methods. In the *Boruta* method, the feature with the highest importance is *Thallium*, while for the other two methods, it is *Sex* and *Cholesterol* respectively. Additionally, *Number of vessels fluro* consistently ranks among the top 5 most important features across all methods. Consequently, the next step in our research methodology involves exploring the genuine causal relationship between heart disease detection and highly correlated features such as *Thallium, Sex, Cholesterol*, and *Number of vessels fluro*.

**Fig 1.**
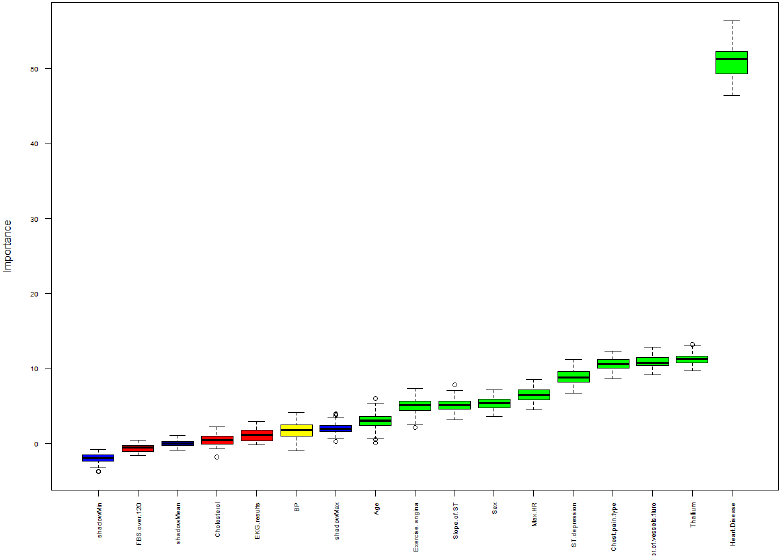
The distribution of importance scores for each feature based on the *Boruta* method, from the least important features to the most important one (left to right)

**Fig 2.**
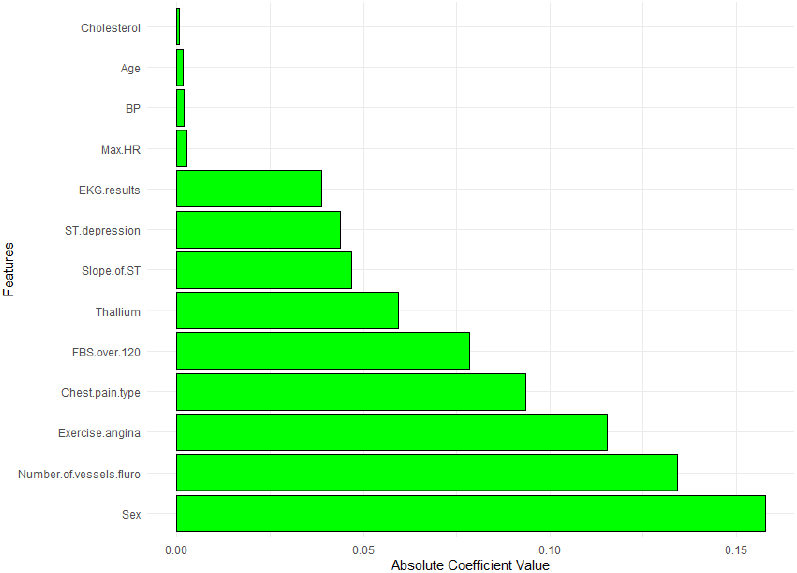
Absolute coefficient values of selected features in the Lasso Regression method, ordered from the least important features to the most important one (top to bottom)

**Fig 3.**
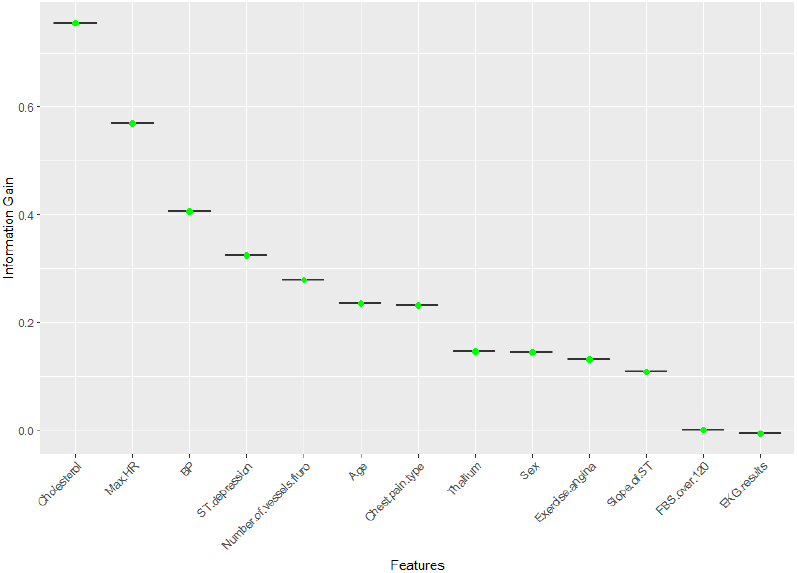
The distribution of Information Gain (IG) scores for each feature in the Information Gain method, ordered from the most important to the least important (left to right)

In causal analysis, examination involves the consideration of a treatment variable, an outcome variable, and various confounding variables. In the context of the dataset under consideration, the treatment variable is one in the subset *S* ={*Number of vessels fluro, Thallium, Sex, Cholesterol*}. The outcome variable is identified as the presence or absence of heart disease, while confounding variables encompass subset *S* minus the treatment variable.

Regarding the method of analysis, we are restricted to binary treatment and outcome variables. The first treatment considered is *Number of vessels fluro* which shows the value for the numbers of vessels shown in the Fluroscopy images. This variable in the initial dataset has four values of {0, 1, 2, 3}. To make this treatment variable into a binary treatment variable, a new metric is defined: If the variable has the value of 3, it;s binary counterpart would be equal to 1 otherwise it would be zero. Therefore the new *Number of vessels fluro* is:

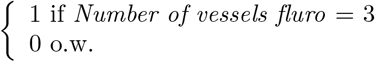

The second treatment variable is Thallium with values {3, 6, 7}. Value 3 for Thallium shows a normal thallium test while value 6 shows defect but not ischemia which refers to a condition where there is a reduced blood flow to a part of the body, typically due to a blockage or narrowing of the blood vessels. However value 7 is a strong indicative of ischemia. So in order to make this treatment variable binary we assign 1 to value 7 and otherwise 0. The new treatment variable Thallium is defined as:

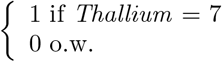

The third treatment variable which is Sex is already a binary treatment variable with 0 being assigned to Female and 1 assigned to male patients. The last treatment variable in the subset S is Cholesterol. Here cholesterol is a continuous integer value with minimum of 126 and maximum of 564 with the mean of 249.65. To make this binary, we define the threshold of 240 as being high indicated by 1 and otherwise 0 as advised by CDC guidelines. The new treatment variable is:

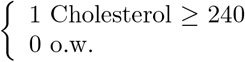

Also the outcome variable is defined as below:

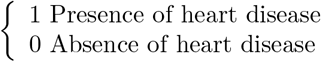

Utilizing the G-computation method, the causal relationship between an individual’s Thallium level, Number of vessels shown in Fluroscopy images, their gender or cholesterol levels and the likelihood of developing heart disease was assessed. The G-computation method is a recognized approach that enables the identification of causal relationships by incorporating counterfactual notions. It involves considering different scenarios and ‘what if’ situations under various treatment regimens, allowing for a comprehensive evaluation of the causal connection between the mentioned predictor variables and the probability of heart disease. Let *T* be the treatment of interest, where *T* =1 when the patient is treated or exposed and 0 otherwise. We can also define the outcome variable as *Y*, with two possible values: 1 (presence of heart disease) and 0 (absence of heart disease). For the confounding variables, also known as covariates, we define the set of variables *C*. The G-computation process is as follows [10]:

#### Step 1: Fitting a Q-model

The process starts with fitting a Q-model which is a logistic regression and can be defined as logit{P(Y=1|T,C)} = *α*T + *β*C [8].

#### Step 2: Calculation of expected probabilities

Then, the expected probability of events under treatment and no treatment can be calculated. For the treated group, the formulation would be 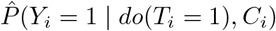, and for the untreated group, the formulation is 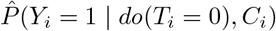. With computing the 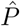 values for all subjects, we get two vectors of probabilities referred to as 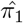 and 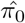.

#### Step 3: Calculating the ATE

ATE corresponds to Average Treatment Effect on the Entire population and calculates the marginal effect if the entire sample is treated versus the case where the entire sample is untreated. One can calculate ATE using the formula

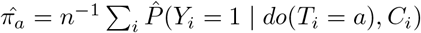

To calculate the variance of our estimator, we can employ either simulation or the bootstrap re-sampling method. Simulation involves simulating the parameters of the multi-variable logistic regression under the assumption of a multi-normal distribution. It has been suggested that both methods can yield similar results, as discussed by [9]. However, one drawback of bootstrap re-sampling is its computational time and the corresponding resource requirements. Since the dataset used in this research is rather small, we have chosen the bootstrap method.

The procedural steps outlined in the previous statement were executed through the utilization of the *RISCA (1*.*0*.*4)* package within the *R* programming language. The comprehensive analysis of the outcomes was also conducted within the *R* environment. Specifically, the function employed from the *RISCA* package for these tasks was identified as *gc*.*logistic*, as documented in [26]. The computational analyses for this study were conducted using a personal computer as the primary computing resource with an *Intel* 13th Generation Core i7 processor, 16 gigabytes of RAM, and 512 gigabytes of storage capacity.

## 2 Results

The results of the G-computation can be observed in Table 4. The first sub -table displays the values for 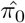, which represents the average proportion of events in the unexposed/untreated sample and is calculated as 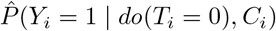 for each treatment variable. In this table, ‘estimate’ denotes the estimated value of 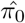, while *ci*.*lower* and *ci*.*upper* represent the 95 percent confidence interval. The second sub-table presents the values for 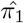, focusing on the average proportion of events in the exposed/treated sample, calculated as 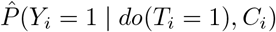.

**Table 4.** Results of the Analysis.

| Original Estimates |  |  |  |  | logOR Estimates |  |  |  |
| --- | --- | --- | --- | --- | --- | --- | --- | --- |
|  | Estimate | ci.lower | ci.upper | std.error | Estimate | ci.lower | ci.upper | std.error |
| Thalium | 0.4495 | 0.0613 | 0.3292 | 0.5698 | 1.9382 | 0.3141 | 1.3225 | 2.5540 |
| Sex | 0.1542 | 0.0612 | 0.0342 | 0.2742 | 0.6243 | 0.2584 | 0.1178 | 1.1309 |
| Vessel | 0.2790 | 0.1051 | 0.0736 | 0.4859 | 1.4234 | 3.6496 | -5.7296 | 8.5766 |
| Cholesterol | 0.1623 | 0.0492 | 0.0659 | 0.2588 | 0.6694 | 0.2098 | 0.2581 | 1.0807 |

Note: Results include original estimates and logOR estimates with standard errors.

The third sub-table is dedicated to the difference between the average proportions of events in the exposed/treated sample and the unexposed/untreated sample, referred to as *delta*. In this table, *std*.*error* represents the corresponding standard error. Finally the last sub-table in Table 4 represents the logarithm of the average Odds Ratio (OR).

The final output of *gc*.*logistic* function and the most important finding that answers our main question is revealed is *p*.*value* which is shown in Table 5. As the name indicates, the p-value is corresponding to the bilateral test of the null hypothesis *π*_0_ = *π*_1_, i.e. OR = 1, meaning there is no causal relation between the treatment and the outcome.

**Table 5.** P-value and ATE results for G-computation analysis.

| Variable | P-value | ATE |
| --- | --- | --- |
| Number of vessels fluro | 0.6773 | 0.2798 |
| Thallium | 3.2708e-10 | 0.4495 |
| Sex | 0.0182 | 0.1542 |
| Cholesterol | 0.0011 | 0.1623 |

Table notes: Results show the P-value and ATE for different variables in G-computation analysis.

In general if p-value is insignificant, usually smaller than 0.05, this means that the null hypothesis can be rejected.

To analyze the causal relation between Thallium stress test results and the detection of heart disease, we first examine the p-value, which is nearly zero. This suggests that we can reject the null hypothesis, indicating no causal relation between the results of the Thallium test and heart disease. Thus, even under different scenarios involving varying sexes, cholesterol levels, and the number of vessels shown in fluoroscopy, the stress test results can significantly indicate a causative path to heart disease. The next question this research aims to answer is: What is the average change in the probability of heart disease for individuals who have ischemia, with Thallium test results of 7, compared to those who do not, considering the influence of other covariates included in the model? This probability is represented by the column ATE in Table 5. For the treatment variable, Thallium, this value is 0.4495, indicating that, on average, there is a 45% increase in the chance of heart disease in the presence of all confounding variables, if the patient’s Thallium test results show ischemia or reduced blood flow through the veins.

The p-value from the G-computation analysis regarding cholesterol is 0.001, signifying a clear causal link between elevated cholesterol levels and the risk of heart disease. Examining the Average Treatment Effect (ATE), we find that, holding all other confounding variables constant (e.g., for two women with normal thallium test and fluoroscopy images), an individual with cholesterol levels of 240 or higher has, on average, a 16% greater likelihood of developing heart disease compared to those with lower cholesterol levels. The results suggest that managing cholesterol levels can be considered as a preventive measure against heart disease.

By looking at the findings of Table 5, it can be observed that heart disease is not gender-neutral. The data suggests a significant gender disparity in the likelihood of experiencing heart disease, even when key health indicators such as cholesterol levels, fluoroscopy images, and Thallium test results are identical between female and male participants. Specifically, the analysis reveals that there is still a 15% chance that a male patient will develop heart disease while a female patient with the same health profile will not. This indicates that factors beyond the traditional risk markers, possibly including genetic, hormonal, or lifestyle differences, may play a role in the increased susceptibility of men to heart disease.

These results highlight the importance of gender-specific approaches in public health strategies aimed at combating heart disease. Policymakers can leverage this information to develop targeted interventions that address the unique needs of men. For instance, special programs could focus on encouraging regular medical check-ups, promoting heart-healthy diets, and raising awareness about the specific symptoms of heart disease that men might experience. By tailoring these initiatives to the male population, it is possible to mitigate the higher risk observed in men, ultimately reducing the incidence of heart disease and improving overall public health outcomes.

Until now, all the features with the highest correlation in different feature selection methods also have some significant causal relation to the detection of heart disease. So does this mean correlation equals causation? By looking at the results of fluoroscopy images, we can see that there is no causal correlation between the amount of blocked veins in the patient’s body and the detection of heart disease since the null hypothesis cannot be rejected because p-value has a significant value of 0.67. This means that even though in all three methods of feature selection, fluoroscopy images were of high importance for accurate prediction of heart disease presence, this does not indicate that by looking at an image of blocked veins, we can easily infer that the mentioned patient will have a heart disease condition and the blocked veins caused the heart disease in the patient.

Table 6 shows the estimates of the ORs in tree cases. The first case shown as *Raw*, depicts the raw effect of the treatment which is defined as a simple logistic regression of treatment and outcome. *Conditional* shows the value of logistic regression in the case where our three covariates are considered as well. The first two values only focus on the simple and advance correlation of the treatment and outcome. But the *Marginal* value shows the causal association of treatment and outcome where all the counterfactuals and “what-if”s were considered to see the effect of treatment on the outcome in the presence of the confounding variables that impact both the treatment and outcome.

**Table 6.**
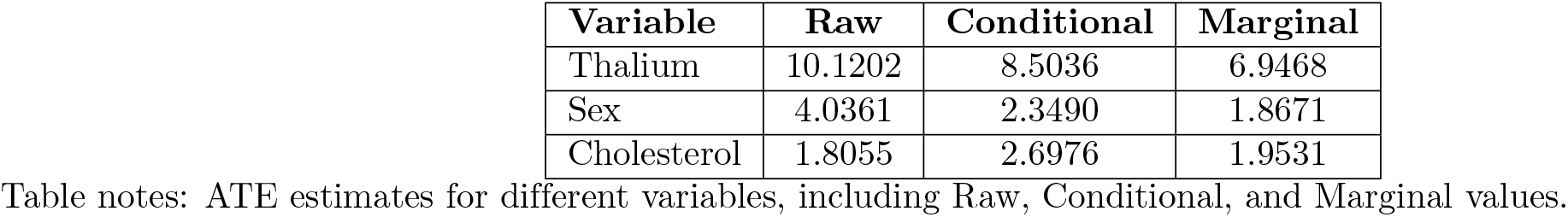
ATE Estimates.

For the case where the treatment of interest is Thallium test the number 10.1202 shows that when considering no other confounding variables, the odd of having a heart disease is approximately ten times higher if the results of the Thallium test show some blockage meaning the results of Thallium test appears to significantly increase the odds of heart disease in this simple model. However when considering other factors such as cholesterol and the sex of the patient the probability of predicting a heart disease while looking at thallium test results this probability diminish to a lower value of 8.5036 but even after accounting for other factors, high cholesterol remains a strong predictor of increased odds of heart disease. The last number which is Marginal Effect (6.9468) represents the Average Treatment Effect (ATE), considering the propensity score and possibly other adjustment methods (gc-logOR). On average across different scenarios (different levels of Sex, Fluoroscopy test results and cholesterol), the odds of heart disease are about 6.9468 times higher in individuals with high Thallium results compared to those without that. All three odds ratios (raw, conditional, and marginal) indicate a significant association between high cholesterol levels (your treatment variable) and increased odds of heart disease. The decreasing values from raw to marginal suggest that as you account for more variables or use different adjustment methods, the magnitude of the effect slightly decreases, but it remains substantial and statistically significant across all models

This also holds true for the other two treatments considered however the odds are lower. Meaning that the probability of having a heart disease if for male patients or patients with high cholesterol levels while considering other confounding variables are approximately two times higher than the probability of having a heart disease problem for a female patient of a patient with low cholesterol levels. But this odds are not as strong as the case of Thallium test results. This suggests that investing on policies that help reducing the blockage of vein in a patients body has a much more higher impact on preventing heart disease.

## 3 Discussion & Future Research

The result of this research proposes that while usual machine learning techniques, which use predictor variables such as a patient’s sex and health levels, can to some extent provide a good understanding of the outcome, these predictions mainly show a high correlation between the outcomes and the predictor variables. However, if the goal is to give personalized advice to individuals, as advised by healthcare practitioners, or to set up general healthcare policies established by governments, these correlations may not be sufficient. This research first uses highly demanded techniques in machine learning to detect the most important features. Then to delve into the causal relation of those variables by using the g-estimation technique. The results show that while all the features selected by feature selection algorithms are showing high correlation between Cholesterol, sex, fluroscopy results and Thallium test, not all of them are the cause of heart disease.

For the results of thallium test we can detect a high very significant causation between the reduced flow of blood in vessels due to narrowing of vessels or blockage. This shows that on the individual levels healthcare providers such as doctors can prescribes Vasodilators. Vasodilators are a group of medicines that dilate open blood vessels and keeps them from contracting. Also since a healthy diet is a primary part of a healthy heart the results shows that advising patients with a family risk of heart disease to choose diets implemented with vitamins E and B ccan help them with blood flow. On the higher level governments can develop practices where public exercise is advised since the best practice to enlarge the veins is strength based exercises. They can assign or facilitate gyms with strength workout.Government can also invest in public infrastructure like parks, walking and biking trails, and recreational facilities to encourage physical activity. Organize community fitness programs and events.

Another strong causation proven by this research is sex of the patients. Meaning that when two patients with the same thallium test results and cholesterol levels, the male patient is more likely to develop heart disease. So, this research can be used by governments and public health care providers to inform males from a young age that they are more likely prone to heart failures so for them keeping track of their healthy habits starting from a young age is much more important that others.

The other significant causation found was between cholesterol levels and the detection of heart disease. While until now there have been many practices encouraging people to adopt a healthier lifestyle, this research shows that it can be really effective. Policy makers can launch public awareness campaigns to inform the public about the dangers of high cholesterol and the benefits of a healthy lifestyle. They can also add clear and comprehensive nutritional labeling on food products, highlighting cholesterol content and encouraging healthier choices. Another option is providing subsidies for fruits, vegetables, whole grains, and other heart-healthy foods that directly affect cholesterol levels and implement taxes on high-cholesterol and high-fat foods to discourage consumption.

However although some factor causing heart disease failure were detected by this research also suggests that not all correlations means causation. For example for the case of fluoroscopy results, not all blocked veins represent a definite cause leading to heart disease. Meaning that when planing for improving heart health, those results may not be important enough and they cannot be the solely metric which we plan the heart disease on. This finding can help doctors to account for other metrics first when they plan to evaluate heart disease of a patient.

In this research, a dataset with 13 predictor variables was used to evaluate the detection of heart disease. Among those 13 predictor variables, 4 of the most important features were selected using well-known feature selection methods. For future research, more feature selection methods can be tested, and more tailored methods can be constructed to work best on this dataset. Additionally, more confounding variables, such as smoking and a history of heart disease, can be considered so the effect of those treatments can be more thoroughly investigated. In this research, for analyzing the causation between the treatment variables and the outcome, which is the presence of heart disease, the well-known method of g-estimation was used. However, more sophisticated methods of causal inference can be used to better detect the causal hierarchy as well.

## 4 Conclusion

In this research, our aim was to detect a causal relationship, not just a correlation, between some important health factors in a patient, such as sex, cholesterol levels, blocked veins, and blood flow, and the detection of heart disease. To achieve this goal, three well-known feature selection methods were considered. The most important features identified by these three methods, including one feature that was consistently important across all three, were selected. Then, causal analysis was carried out using the well-established method of G-estimation to calculate the average marginal effect of treatment on the entire population. The results once again restate the fact that correlation does not imply causation. The difference between using machine learning techniques and conducting a causal analysis is that in the latter, one is not confined to fitting a regression model on two variables but is dedicated to exploring all possible outcomes that could have arisen with different treatment regimens. While all the treatment variables showed a high correlation with the outcome variable, only three—cholesterol levels, sex of the patient, and blood flow pattern in their vessels—were found to significantly increase the odds of heart failure when considering other variables. The proven causation between sex, cholesterol levels, and blood flow in patients can help healthcare practitioners and policymakers focus more on these factors, such as advising the consumption of healthy foods like whole grains and fruits and vegetables and implementing policies that promote exercise habits to impact heart disease.

## Data Availability

All CSV files are available from the Kaggle database (URL: https://www.kaggle.com/datasets/fedesoriano/heart-failure-prediction).

## Notes

### Competing Interest Statement

The authors have declared no competing interest.

### Funding Statement

The author(s) received no specific funding for this work.

